# Safety and efficacy of orally administered full-spectrum medicinal cannabis plant extract 0.08% THC (NTI-164) in children with autism spectrum disorder: An open-label study

**DOI:** 10.1101/2023.12.05.23299505

**Authors:** Dima El-Sukkari, Kanan Sharma, Bobbi Fleiss, Dion L. Braganza, Alison Crichton, Michael C. Fahey

**Affiliations:** Monash Children’s Hospital, Clayton, Victoria, Australia; RMIT University, STEM College, Bundoora, Victoria, Australia; Department of Paediatrics, Monash University; Developmental Paediatrics, Department of Paediatrics, Monash Children’s Hospital

## Abstract

Autism spectrum disorder (ASD) is commonly associated with debilitating comorbidities impacting the well-being of affected children and their families. Some children with ASD experience behavioural difficulties that do not respond well to current medications and may also experience unwanted side effects. Therefore, it is crucial to develop alternative, safe and effective therapies. The improved understanding of the endocannabinoid system, together with emerging evidence for the therapeutic effects of cannabis derivatives in neurodevelopment disorders, has led to an exploration of their use in ASD.

This open-label study assessed the efficacy and safety of a novel oil-based full-spectrum medicinal cannabis plant extract 0.08% THC (NTI-164) in treating 14 children with ASD symptoms (13.4 years, range 10-17). Data on the safety profile of NTI-164 was collected through biochemical analysis, vital signs, and parent and participant reports. The efficacy was assessed through a dose-escalation protocol using a broad range of validated clinical behavioural assessments and parent and child-reported questionnaires.

Following four weeks of treatment with NTI-164, 93% of participants demonstrated significant overall improvement in ASD-related symptoms compared to baseline with transient side effects that did not interfere with their general functioning. In addition, targeted behavioural problems were rated as much improved or very much improved in 46% of the treated patients. More than half of caregivers and children also reported decreased anxiety symptoms.

The findings from this study suggest that NTI-164 is well-tolerated and safe, with potential clinical benefits in improving disruptive behaviours and reducing anxiety in children with ASD-related symptoms. Future longitudinal and well-controlled studies are warranted to develop evidence-based clinical therapies and further evaluate the therapeutic benefit of full- spectrum cannabis extracts in managing ASD core and associated comorbid symptoms in this group of children.

**Trial registration number** ACTRN12621000760875

## Introduction

Autism Spectrum Disorder (ASD) is a group of pervasive neurodevelopmental disorders that appear in early childhood. These disorders are characterised by social functioning deficits, restricted or repeated patterns of behaviour across the lifespan [1], and other comorbidities such as intellectual disabilities, epilepsy, anxiety, hyperactivity, irritability, and sleep disturbances. [2, 3] These symptoms can significantly impact a person’s daily functioning. [4, 5] Globally, the prevalence of ASD is estimated to range from 0.4% to 1.7% [6]. In the last decade, the prevalence rate of young children (under five years) diagnosed with ASD has increased by 6.7%, from 2.7 million in 1990 to 2.9 million in 2019 [7].

The families of children with ASD face a substantial financial burden, with recent global estimates indicating 2834 EUR total costs for health-related services combined with indirect societal costs over two months [8]. The estimated lifetime social cost in the US alone was $7 trillion for the last three decades (1990-2019) and is forecasted to increase to $11-15 trillion in the following decade [9]. The continuing rise in ASD prevalence is further projected to generate an enormous economic burden on US healthcare for decades and to reach an annual cost estimate of $5.54 trillion by 2060, with substantial potential yearly savings of $1.9 trillion through ASD prevention strategies [10]. Despite thorough investigation, no evidence supports the effectiveness of pharmacological treatments in relieving some of the symptoms that cause people with ASD distress. [11, 12]. However, these pharmacological treatments often result in adverse side effects in children, with a notable variation in effectiveness between individuals [13] and a lack of adherence [14]. The complexity of the ASD behavioural phenotype and its aetiology resulting from the interaction of multiple genetic and environmental risk factors further limits targeted interventions and the development of effective therapies [15, 16]. Therefore, to address core behaviours and co-morbid symptoms associated with ASD, targeted therapeutic interventions are urgently needed.

The endocannabinoid system (ECS), consisting of cannabinoid receptors (CB1 and CB2), endocannabinoids (endogenous bioactive ligands) and metabolic enzymes [17] presents as a potential pharmacological target for developing therapeutics in the management of ASD symptoms since it has a crucial role in modulation of neuronal plasticity during development [18] and associated with social-emotional processes [19] which are impaired in ASD-related phenotypes [20, 21]. The central endocannabinoid receptors CB1 and CB2 are expressed mainly in sensory neurons and immune cells, including microglia [17]. The endogenous cannabinoids are produced as required, activating presynaptic receptors to reduce calcium uptake and inhibit the release of neurotransmitters, consequently influencing a broad range of biological responses [22]. Alterations in ECS functionality have been reported in several preclinical ASD models [23–25] and clinical studies [26–28], suggesting that ASD pathogenesis likely involves dysregulation of the endocannabinoid signalling pathway. These findings suggest that endocannabinoid anandamide (AEA) and its associated metabolic enzymes may be a potential therapeutic target for ASD-related behavioural impairment. In animal studies, treatment with inhibitors of the enzyme fatty acid amide hydrolase that degrades anandamide resulted in increased AEA levels, restoring cognitive and social deficits in the ASD-related knockout models [29–31].

The increased understanding of the ECS signalling concurrent with growing evidence for the therapeutic effects of cannabis (*Cannabis sativa* L.) in neurodevelopment disorders has been the catalyst for extensive research on the two most abundant active constituents Δ9-Tetrahydrocannabinol (THC) and Cannabidiol (CBD). During the last two decades, there has been a surge in clinical trials of CBD owing to the absence of psychoactive effects and its confirmed favourable safety profile in humans for the treatment of several neurological and psychiatric disorders [32–34]. In addition, accumulating evidence from experimental and animal model studies on the effect of CBD has demonstrated modulation of a broad range of biological responses, including antidepressant [35], anti-inflammatory [36], antinociceptive [37], anti-seizure [38], immunomodulatory [39] and neuroprotective activities [40]. Cannabinoids are generally well-tolerated, but evidence suggests that the risk of hepatotoxicity may be increased when combined with other medications. This risk is usually low, dose-dependent, and more likely to occur in individuals with a history of liver disease.[41] Rigorous controlled clinical studies are needed to determine the efficacy and safety profile of cannabinoids in children.

There is growing evidence that using CBD in combination with other plant compounds found in cannabis as a full-spectrum botanical medicine could provide unique therapeutic benefits, as suggested by anecdotal reports and emerging research in the field. [42] This effect, known as the entourage effect, was initially proposed as a regulatory mechanism for the endocannabinoid system, whereby multiple compounds enhance the activity of endogenous cannabinoids. A recent meta-analysis of observational studies showed that twice as many patients with treatment-resistant epilepsy improved when treated with an herbal formulation that offered a better safety profile than isolated CBD product [46]. Furthermore, promising evidence from preclinical cancer models supports the potential for the entourage effect of botanical preparations to produce enhanced therapeutic responses than individual cannabinoids [47–49]. Given that the current single molecule approach of individual phytocannabinoids remains the predominant pharmaceutical development strategy [50], for complex diseases such as ASD, multi-target strategies and botanical drug development may be an effective treatment strategy.

Evidence from animal studies suggests that CBD positively affects various physiological responses. However, there is limited evidence from clinical trials to support the potential therapeutic benefits of this non-psychoactive phytocannabinoid for pediatric neurodevelopmental disorders [51]. Only three randomised controlled studies have been published on medicinal cannabis use in children with ASD; two were short-term [58, 59], and the other only reported sleep outcomes [60]. Additionally, most studies on this topic are open-label observational or retrospective design [52–57].

This single centre, phase I/II open-label study aimed to evaluate the efficacy and safety of a novel oil-based full-spectrum medicinal cannabis plant extract with 0.08% THC (NTI-164) in children with ASD over four weeks of daily treatment.

## Materials and methods

### Study design and participants

The study design was a single-centre open-label trial to evaluate the safety and efficacy of a novel full-spectrum medicinal cannabis plant extract containing only 0.08% THC (NTI-164) in children with ASD over four weeks of daily treatment. The trial enrolled 18 eligible male and female paediatric patients aged 8-17 with Level 2 or 3 ASD based on the Autism Diagnostic Observational Schedule (ADOS-2) criteria [61] between 4 February 2022 and 8 September 2022.

This study was approved by the Monash Health Human Research Ethics Committee (RES-21-0000-177A) and registered with the Australian and New Zealand Clinical Trial Registry (Trial ID: ACTRN12621000760875).

### Study objectives

The primary objective was to evaluate the safety and tolerance of NTI-164 in children with ASD throughout the trial. In addition to assessment questionnaires, safety data were collected through laboratory tests, including full blood examination, liver and renal function tests performed at screening and at the beginning of week five during hospital visits. The secondary objective was to assess the efficacy of NTI-164 in treating ASD-associated symptoms. Efficacy data were collected through physician, parent (or caregiver) and school-aged participant-reported questionnaires at two designated time points. This study explored the effect of NTI-164 on core behavioural symptoms of children with ASD and associated symptoms, including irritability, hyperactivity, mood, anxiety, sleep disorders and self-stimulation.

### Pharmacological Intervention

The chemical composition profile of the investigational product NTI-164 consisted of 62% Cannabidiolic Acid (CBDA), 14% Cannabidiol (CBD), 0.44% Cannabigerolic Acid (CBGA), 0.06% Cannabidivarin (CBDV) and 0.08% Tetrahydrocannabinol (THC). The investigational product was provided and dispensed in a glass jar as a 53 mg/mL oil-based solution ready for self-administration at home. During the initial hospital visit (baseline), the participants received the packaged investigational product with instructions. Participants were also given access to a web app (Caspio Participant Portal) to record daily drug administration.

Several lines of evidence from controlled trials have shown the effectiveness of CBD at a dose of 20 mg/kg in managing seizures [62–64]. Therefore, this dose was used in the current study, with participants starting treatment on a daily dose of 5mg/kg of NTI-164 at baseline. To minimise the risk of undesired side effects, the dose was increased by 5mg/kg weekly to reach the maximum tolerated daily dose or 20 mg/kg daily. The daily dose was calculated by multiplying the dosage by each patient’s weight and then dividing by the concentration of CBDA in the oil (53mg/mL). This provided a total daily volume in mL divided into twice daily doses. Protocol adherence was assessed by the accountability of returned investigational product and packaging at the end of the four weeks.

### Outcome measures

#### Primary outcome measure

The primary endpoint was safety and tolerability of NTI-164 treatment as determined by monitoring and assessment through standard laboratory tests of full blood examination, liver and renal function tests, as well as vital signs in addition to physician and parent (or caregiver) reported questionnaires completed at baseline and after completion of 4 weeks of treatment.

Adverse events were assessed and evaluated by designated study staff through discussions with the participant at the scheduled study visit after the end of four weeks, as well as through phone contacts and clinically significant laboratory results. Whenever an adverse event was reported during treatment at home, the Principal Investigator was notified immediately via the online portal. The Principal Investigator or a designated study staff member promptly followed all adverse events. Documentation of adverse events included details of the event, dates and times, severity, duration and possible relationship to the investigational product.

#### Secondary outcome measure

The efficacy assessments included the administration of both quantitative and qualitative ASD-related questionnaires, with the first four scales described below completed by both the caregiver and clinician. The selected questionnaires were specific to various co-morbidities associated with autism spectrum disorder. The purpose was to evaluate the impact of NTI-164 on symptoms associated with autism spectrum disorder. All scales were administered at baseline (week 1) and post-baseline (week 5).

1. Clinical Global Impression of Improvement for the Caregiver (CGI-I-Ca) [65]: This is a 7-point scale measuring symptom change from baseline.
2. Clinical Global Impression of Improvement for the Clinician (CGI-I-C) [65]: This is a 7-point scale measuring symptom change from baseline.
3. Caregiver Global Impression of Change (CGI-C) Target Behaviour [65]: Reflects clinician’s impression of behaviour change on a 7-point scale ranging from 1=not at all to 7=very severe problem.
4. Clinical Global Impression of Change in Attention (CGI-CA) [65]: Reflects clinician’s impression of change in attention on a 7-point scale ranging from 1=not at all to 7=very severe problem.
5. Clinical Global Impression of Severity of Illness (CGI-S) [65]: Reflects clinician’s impression of severity of illness on a 7-point scale ranging from 1=not at all to 7=among the most extremely ill.
6. Anxiety Scale for Children - autism spectrum disorder – Parent Version (ASC-ASD-P) [66]: Parent/Caregiver form was developed to detect anxiety symptoms in youth with ASD. They are composed of four subscales: Performance Anxiety, Uncertainty, Anxious Arousal, and Separation Anxiety. Items are rated on a 4-point scale (0=never and 3=always). Subscales sum to equal a total score.
7. Anxiety Scale for Children - autism spectrum disorder – Child Version (ASC-ASD-C) [66]:: Child form developed to detect symptoms of anxiety in youth with ASD. Composed of the four Subscales: Performance Anxiety, Uncertainty, Anxious Arousal, and Separation Anxiety. Items are rated on a 4-point scale (0=never and 3=always). Subscales sum to equal a total anxiety score, with lower scores indicating a decreased level of anxiousness. The total score ranged from 6 to 55 and 3 to 50 out of a possible maximum score of 72 for the ASC-ASD-P and ASC-ASD-C version, respectively.
8. Sleep Disturbance Scale for Children (SDSC) [67]: Includes the six subscales of Disorders of Initiating and Maintaining Sleep, Sleep Breathing Disorders, Disorders of Arousal, Sleep Wake Transition Disorders, Disorders of Excessive Somnolence, and Sleep Hyperhydrosis. Items are rated on a 5-point scale where 1=never and 5=always (daily). Subscale scores sum to equal a total sleep score, with a higher score reflecting more sleep-related problems.

### Study procedures

The study schedule of procedures and assessments is presented in Table 1. Participants were recruited through Monash Children’s Hospital (MCH) in Melbourne, Australia. Potential participants attended a screening visit for assessment based on study selection criteria (See Supplementary information, File 1) and were provided with the Participant Information Sheet and Consent Form. After obtaining written informed consent from caregivers, eligible participants underwent a physical assessment, including vital signs, anthropometric measurements, full blood examination, liver and renal function tests as part of the screening process. In case of restrained compliance with blood sample collection, analgesia with nitrous oxide was administered by a trained staff member. At this visit, a parent survey was obtained concerning the medical and family history of the participating child, including current attendance at specific programs or health service utilisation.

**Table 1.**
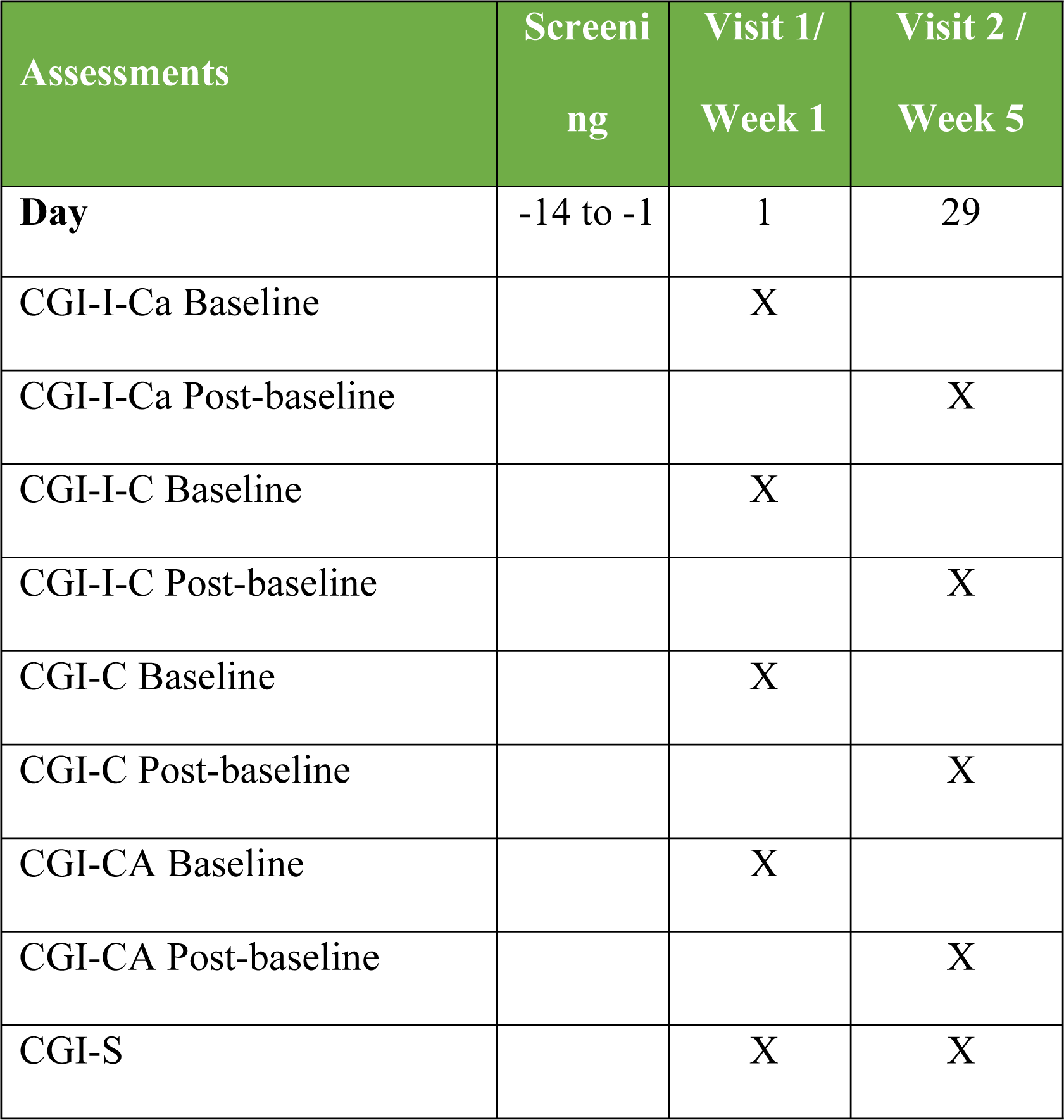
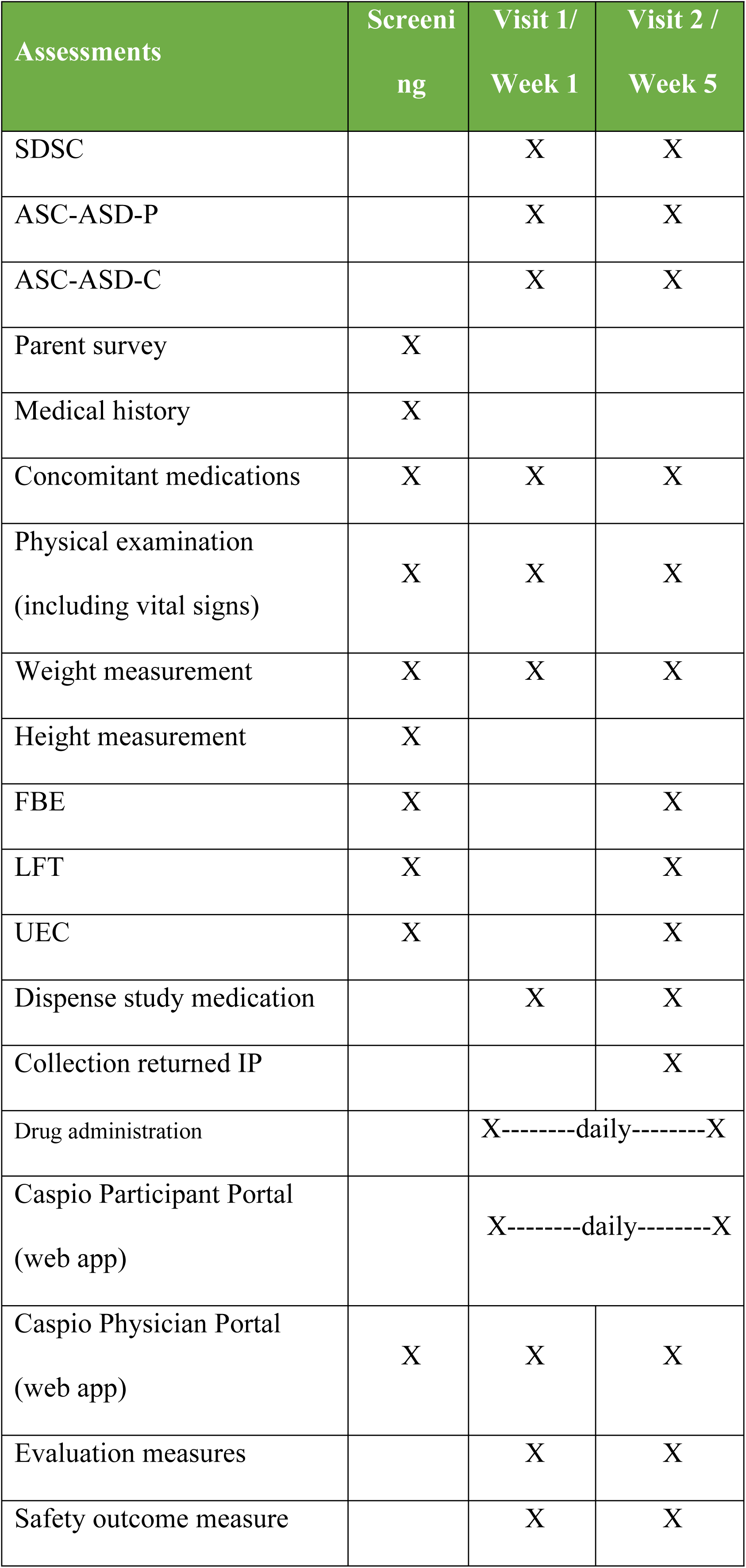
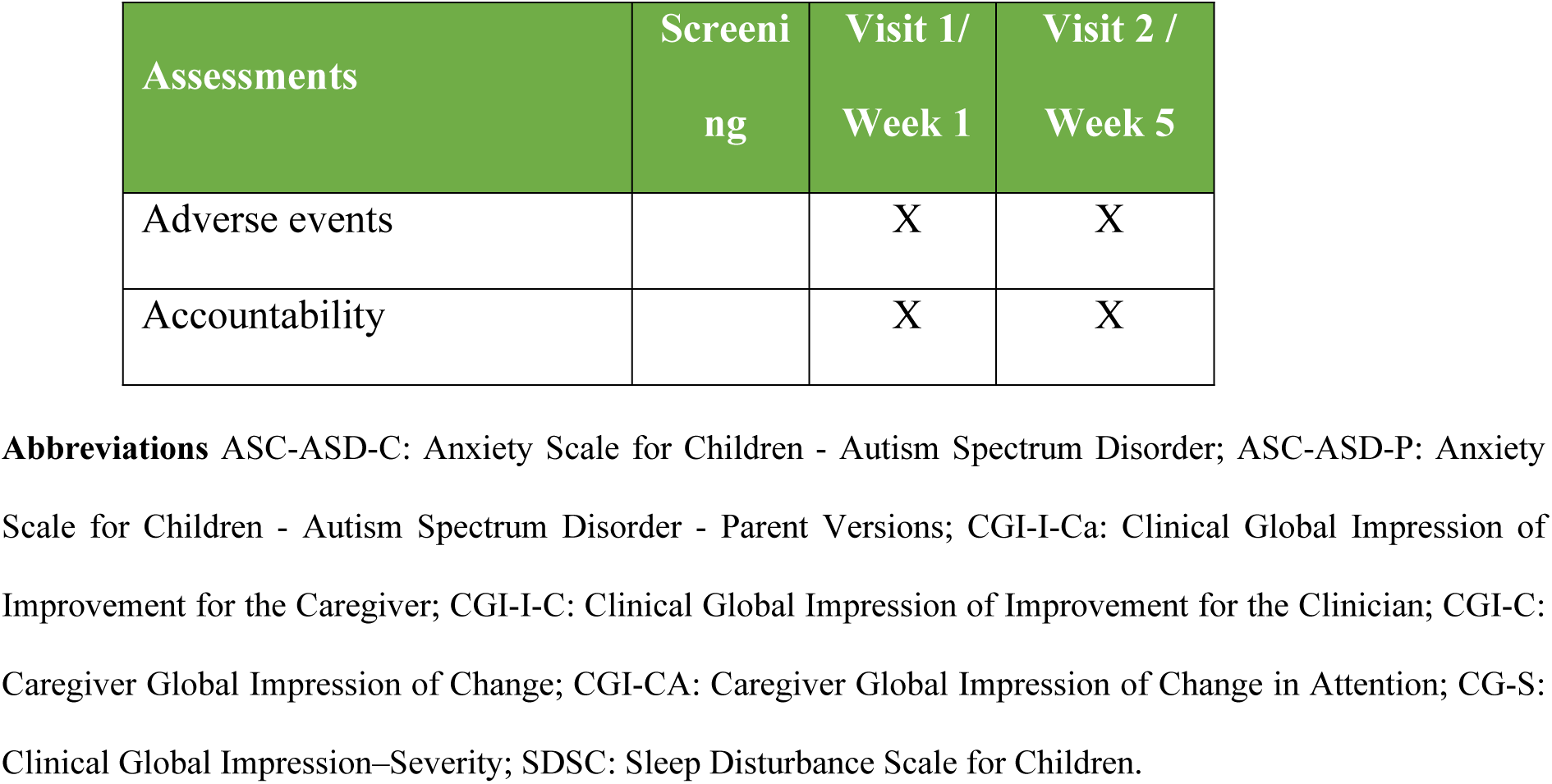
Study schedule of procedures and assessments for the 28 day open-label trial.

| Assessments | Screening | Visit 1/<br>Week 1 | Visit 2 /<br>Week 5 |
| --- | --- | --- | --- |
| **Day** | -14 to -1 | 1 | 29 |
| CGI-I-Ca Baseline |  | X |  |
| CGI-I-Ca Post-baseline |  |  | X |
| CGI-I-C Baseline |  | X |  |
| CGI-I-C Post-baseline |  |  | X |
| CGI-C Baseline |  | X |  |
| CGI-C Post-baseline |  |  | X |
| CGI-CA Baseline |  | X |  |
| CGI-CA Post-baseline |  |  | X |
| CGI-S |  | X | X |

| Assessments | Screeni<br>ng | Visit 1/<br>Week 1 | Visit 2 /<br>Week 5 |
| --- | --- | --- | --- |
| SDSC |  | X | X |
| ASC-ASD-P |  | X | X |
| ASC-ASD-C |  | X | X |
| Parent survey | X |  |  |
| Medical history | X |  |  |
| Concomitant medications | X | X | X |
| Physical examination<br>(including vital signs) | X | X | X |
| Weight measurement | X | X | X |
| Height measurement | X |  |  |
| FBE | X |  | X |
| LFT | X |  | X |
| UEC | X |  | X |
| Dispense study medication |  | X | X |
| Collection returned IP |  |  | X |
| Drug administration |  | X-----daily-----X |  |
| Caspio Participant Portal<br>(web app) |  | X-----daily-----X |  |
| Caspio Physician Portal<br>(web app) | X | X | X |
| Evaluation measures |  | X | X |
| Safety outcome measure |  | X | X |
| Adverse events |  | X | X |
| Accountability |  | X | X |

**Abbreviations** ASC-ASD-C: Anxiety Scale for Children - Autism Spectrum Disorder; ASC-ASD-P: Anxiety Scale for Children - Autism Spectrum Disorder - Parent Versions; CGI-I-Ca: Clinical Global Impression of Improvement for the Caregiver; CGI-I-C: Clinical Global Impression of Improvement for the Clinician; CGI-C: Caregiver Global Impression of Change; CGI-CA: Caregiver Global Impression of Change in Attention; CG-S: Clinical Global Impression–Severity; SDSC: Sleep Disturbance Scale for Children.

At the first visit in week 1 (baseline), participants completed all the required questionnaires and the first daily dose of 5mg/kg of the investigational product NTI-164 was administered. The participant was monitored for 2 hours for any immediate adverse events. Participants who did not report any adverse events were dispensed with NTI-164 for four weeks and instructed to administer the daily volume orally over two doses. Instructions were provided on administering increasing the dose of the product to a maximum of 20mg/kg/day from home. Participants were also trained to use the study web app (Caspio Participant Portal) to record drug administrations, report adverse events, and complete questionnaires from home.

Once the four-week treatment was completed, at the start of the week 5 visit (post-baseline), participants underwent the same set of laboratory tests and physical assessments as before and completed all specified post-baseline questionnaires. To ensure adherence to study protocol and accountability, returned product and packaging were collected and recorded at this visit.

### Statistical analysis

The safety and tolerability were assessed by recording the frequency and severity of reported adverse events by all treated participants. The distribution was analysed using patient demographics, laboratory values and vital signs. Given the small sample size of this study, descriptive statistical analyses of the mean score, mean change, and Standard Deviation (SD) were performed on quantifiable outcome measures. The Wilcoxon Signed-Rank Test and the Paired t-test were used to assess the statistical significance of the analysed data sets. The final statistical analysis included all participants completing the initial four week treatment phase. The CGI-S scale was used to analyse the therapeutic effect of NTI-164 and its changes to the severity of illness. The ASC-ASD-C and ASC-ASD-P questionnaires were used to assess anxiety across four sub-domains, and the SDSC was used to assess changes to sleep from baseline and at the end of the 4 weeks. Clinical Global Impression scales were used to analyse efficacy by measuring symptom change from baseline. Each CGI survey uses the same 7-point scale to measure symptom change from baseline: Very much improved, Much improved, Minimally improved, No change, Minimally worse, Much worse and Very much worse.

## Results

### Participant characteristics

Fourteen of the 18 participants enrolled in the study completed the full treatment. One patient withdrew before treatment began, and three patients discontinued during treatment. Most of the treated patients were male (9/14), Caucasian (12/14), and had an average age of 13.4 years (ranging from 10-17 years). All active patients were diagnosed with ASD Level 2/3 and were evaluated at baseline as “Mildly ill” (4 patients), “Moderately ill” (4 patients), “Markedly ill” (3 patients), or “Severely ill” (3 patients), according to the CGI-Severity of Illness scale. The average daily dose for treated patients was 16.7 mg/kg/day. About 64% of patients could tolerate the maximum dose of 20 mg/kg/day, while 36% tolerated a maximum daily dose ranging between 6 mg/kg/day and 19 mg/kg/day.

### Safety and tolerability

Eight participants experienced ten mild to moderate adverse events, totalling 17 adverse reports during treatment. One participant discontinued treatment. The most frequent side-effect was gastrointestinal symptoms reported by a third of participants (36%). None of these side effects were severe enough to significantly interfere with the patient’s functioning. Most patients were advised to rest to treat these transient adverse effects (Table 2). No serious adverse events or deaths related to the investigational drug were reported during the study. The safety data indicated that NTI-164 at 5, 10, 15 and 20 mg/kg administered in two daily doses is safe and well-tolerated in this study population. Laboratory values provide further safety support. No changes were observed in patients’ full blood examination, liver function or kidney function tests (Table 3). Also, no changes were observed to the vital signs of active patients during treatment, further corroborating that NTI-164 was well-tolerated.

**Table 2.** Reported adverse events during the trial.*

| Adverse Event | Total No. of Reports | Frequency (%) | Treatment |
| --- | --- | --- | --- |
| Abdominal pain | 3 | 14% | Rest |
| Lack/loss of appetite | 3 | 14% | None |
| Diarrhoea | 2 | 9% | None |
| Vomiting | 2 | 9% | None |
| **Lethargy** | 2 | 9% | Rest |
| **Hypoglycaemia** | 1 | 5% | None |
| **Rashes\*\*** | 1 | 5% | Zyrtec for<br>itchiness |
| **Itchiness without**<br>**rash** | 1 | 5% | Warm shower |
| **Sore Throat** | 1 | 5% | Rest |
| **Upper Respiratory**<br>**Tract Infection** | 1 | 5% | Treated by<br>doctor |

\* Two participants were excluded as one had ingrown nail infection and one had knee surgery
\*\* This participant discontinued during the treatment and did not complete the study

**Table 3.**
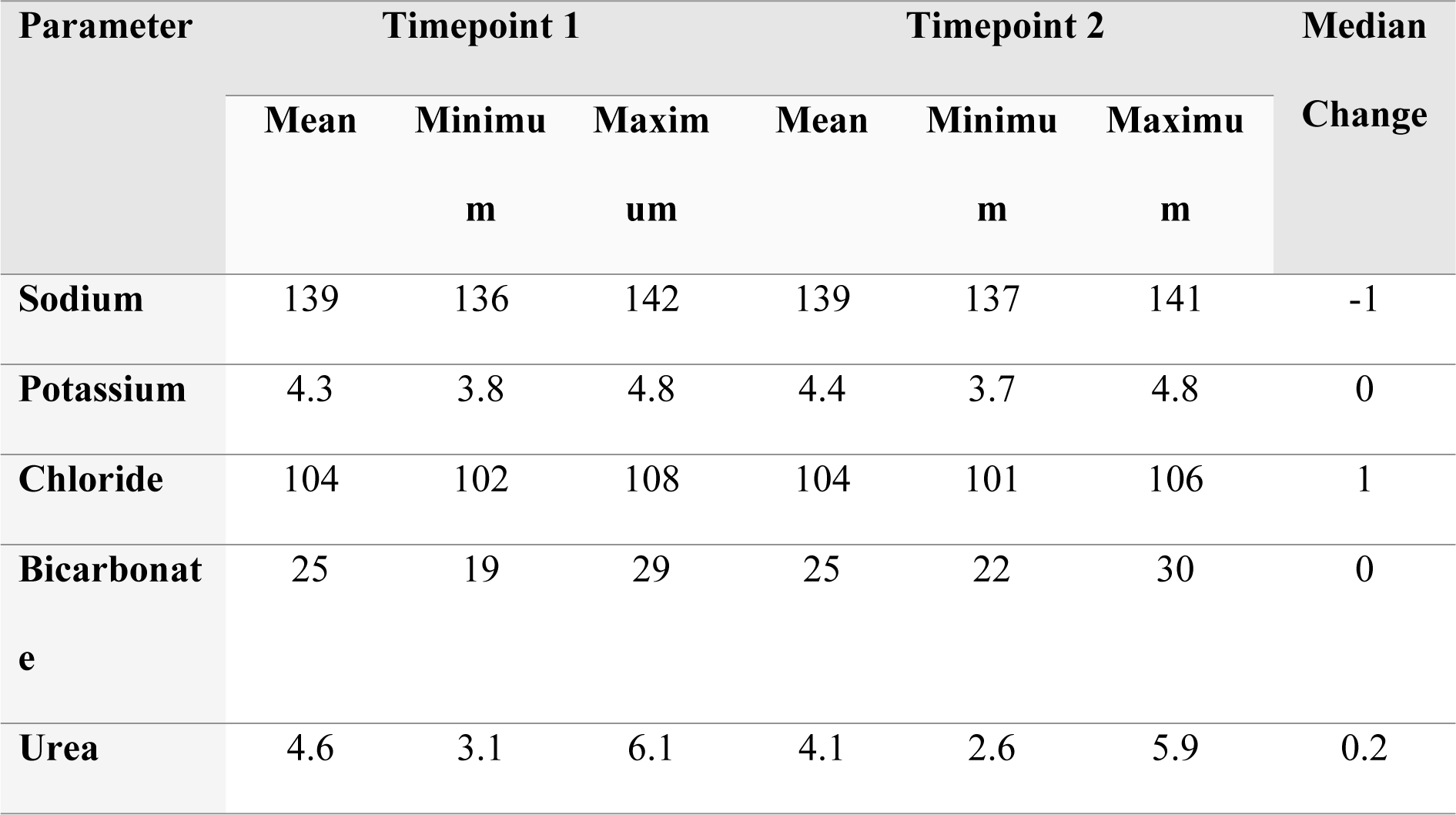
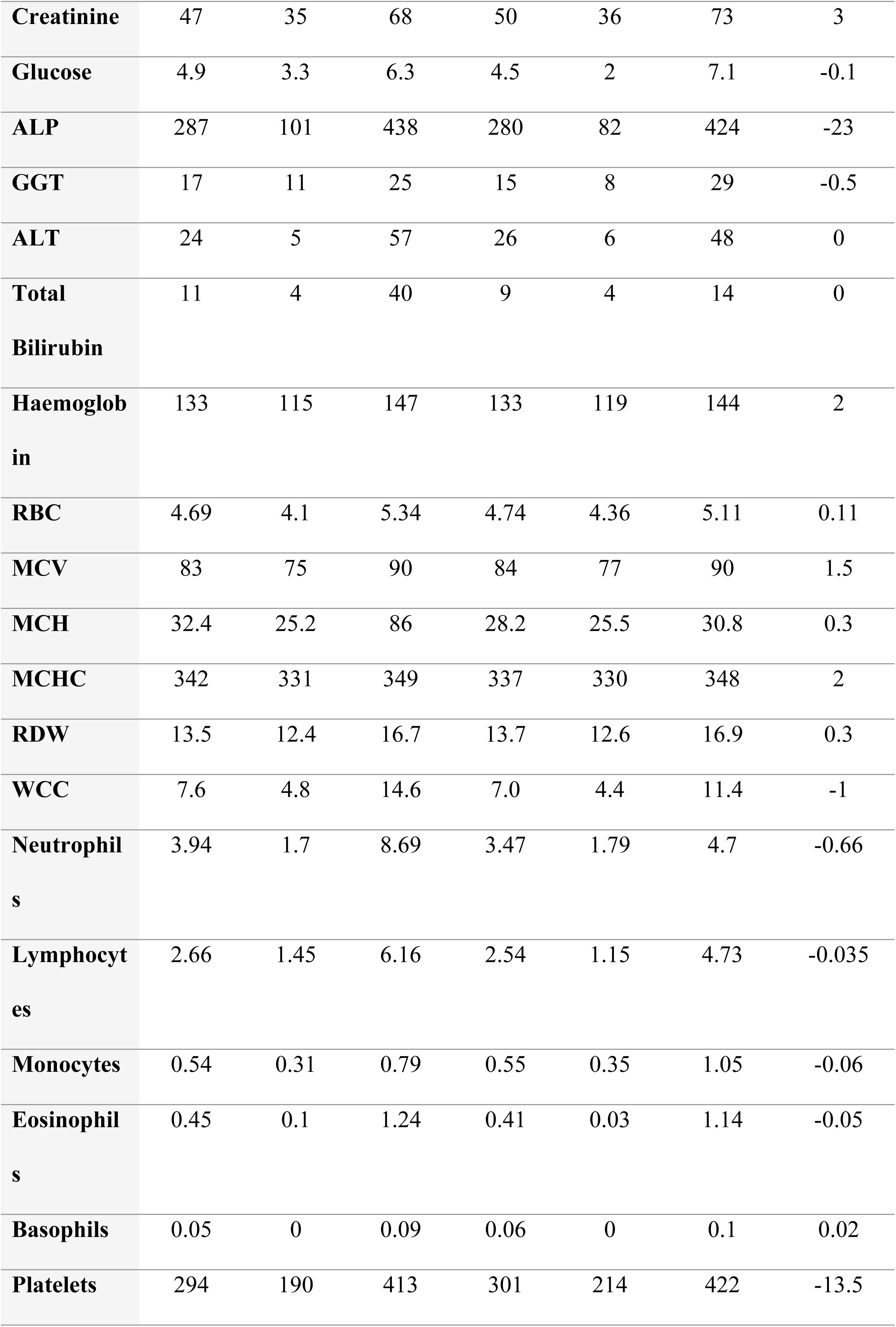
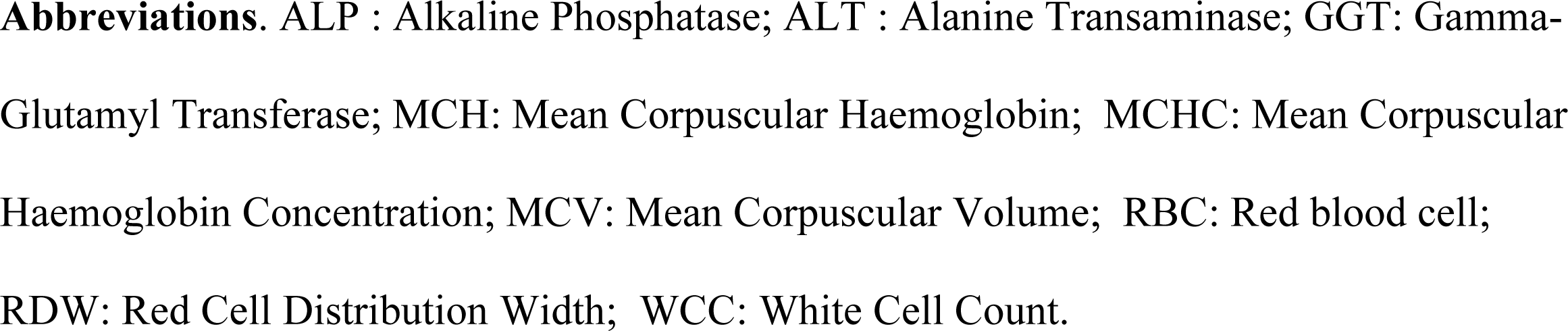
Evaluation of clinical laboratory parameters (n=12) at timepoint 1 (screening visit) and timepoint 2 (start of week 5).

### Analysis of therapeutic efficacy

#### Clinical Global Impression-Severity (CGI-S)

The Clinical Global Impression-Severity (CGI-S) scale was used to assess global improvement, the severity of illness, and the efficacy index based on drug effect only (Table 4). Of the 14 treated patients, 13 (93%) showed symptomatic improvement. 64% of these patients showed a global improvement of ‘Much improved’, 29% showed ‘Minimally improved’, and only one patient (7%) had ‘No change’ (Figure 1).

**Fig 1.**
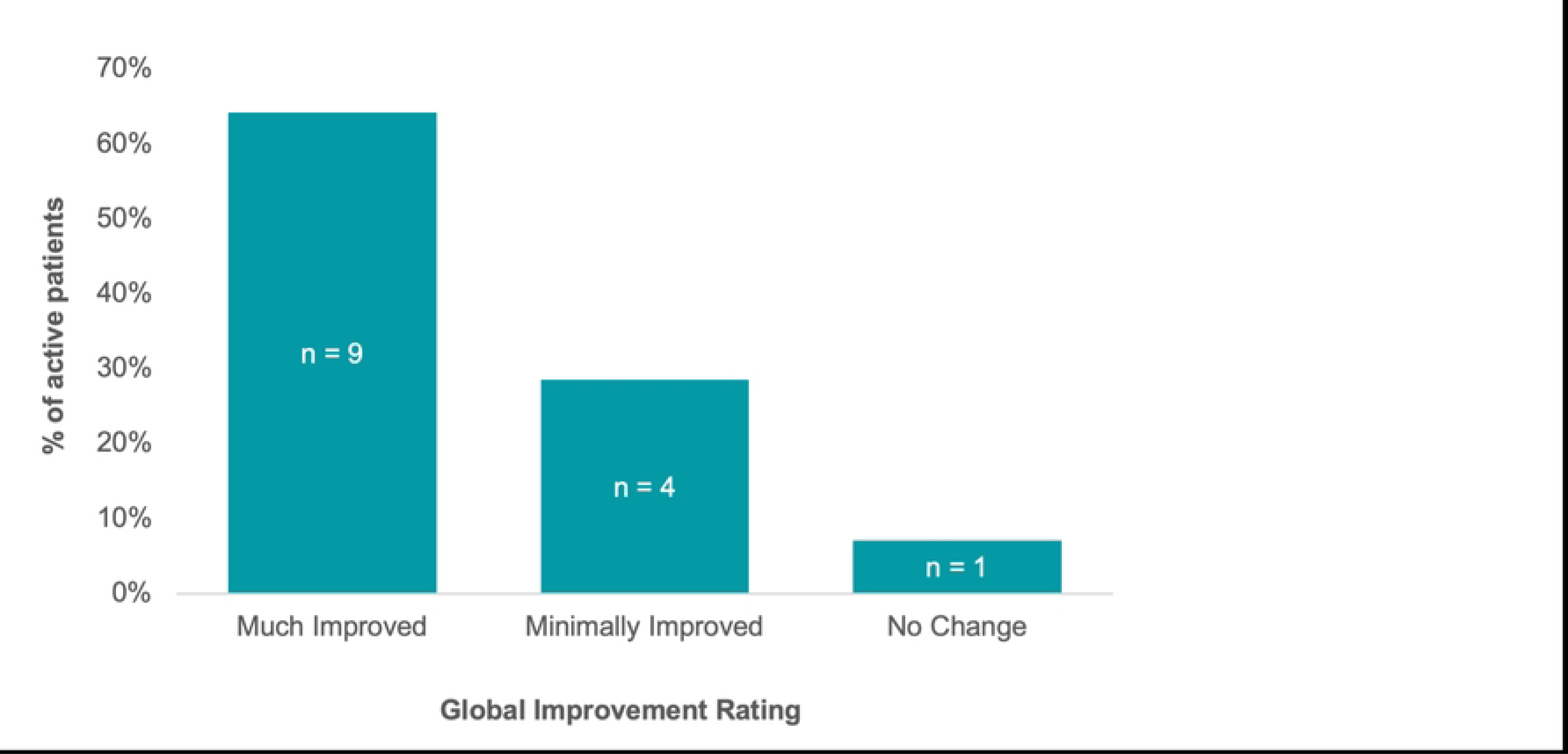
Clinical Global Impression – Global Improvement.

**Table 4.**
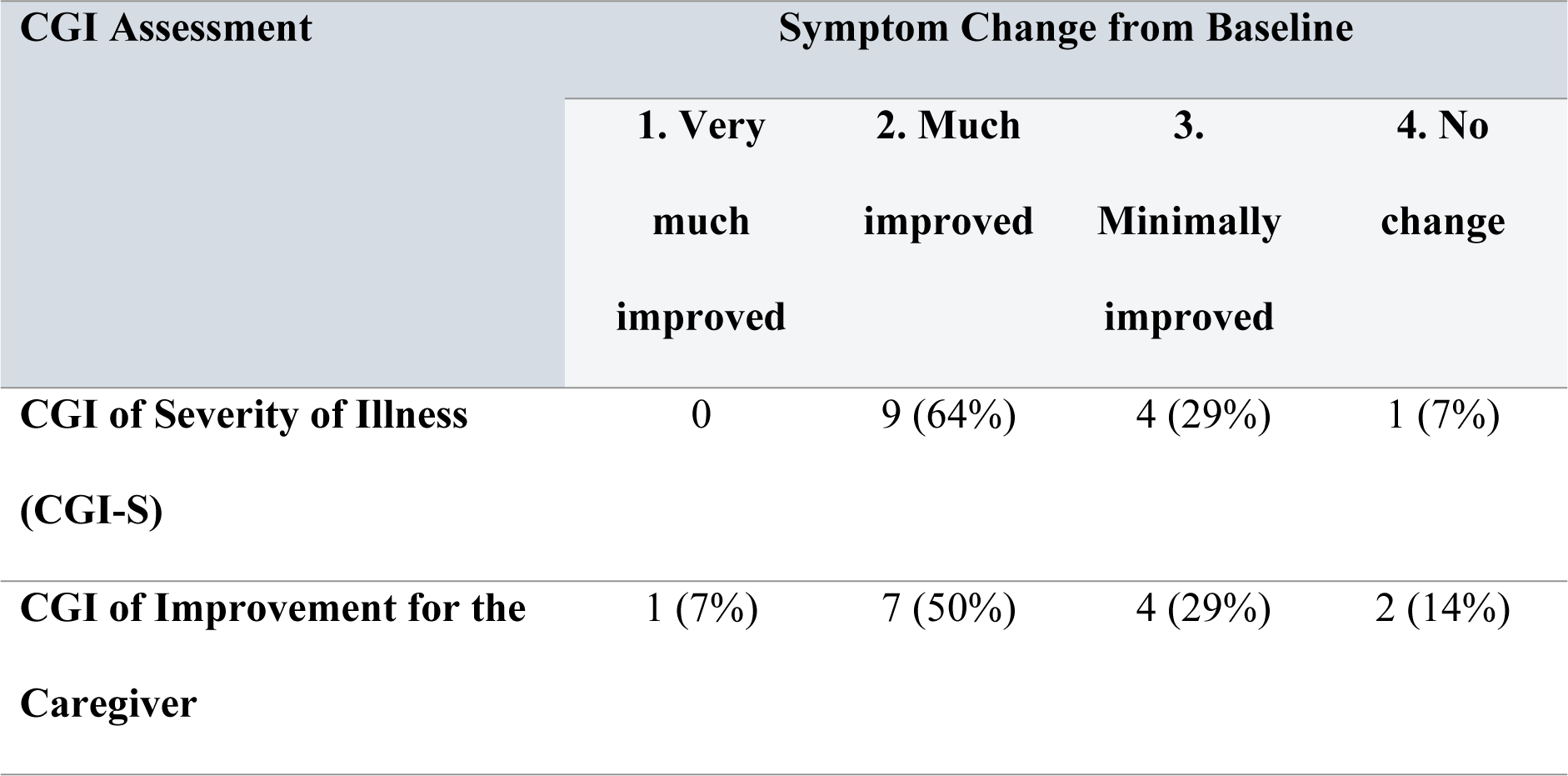
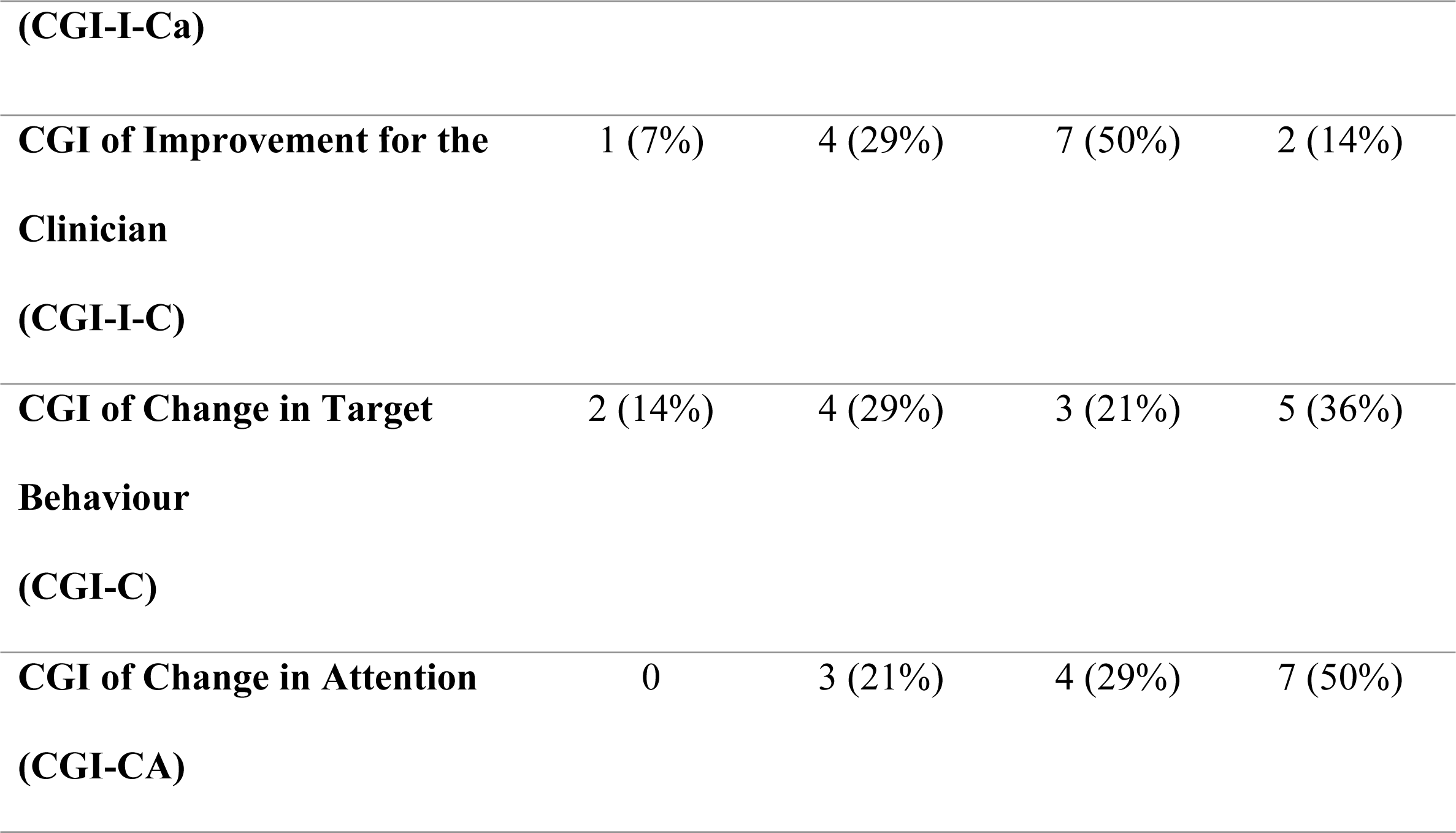
Symptom change scores for each Clinical Global Impression (CGI) scale.

At baseline, the average rating for the severity of illness was 4.4, which was reduced to an average rating of 3.6 after 28 days of NTI-164 treatment (Figure 2). The mean difference of CGI-S between 28 days of treatment and baseline was −0.714 (95% CI: −1.332, −0.097, *p*=0.027), with the Wilcoxon Signed-Rank Test statistic showing a significant difference of −15 (*p*=0.047).

**Fig 2.**
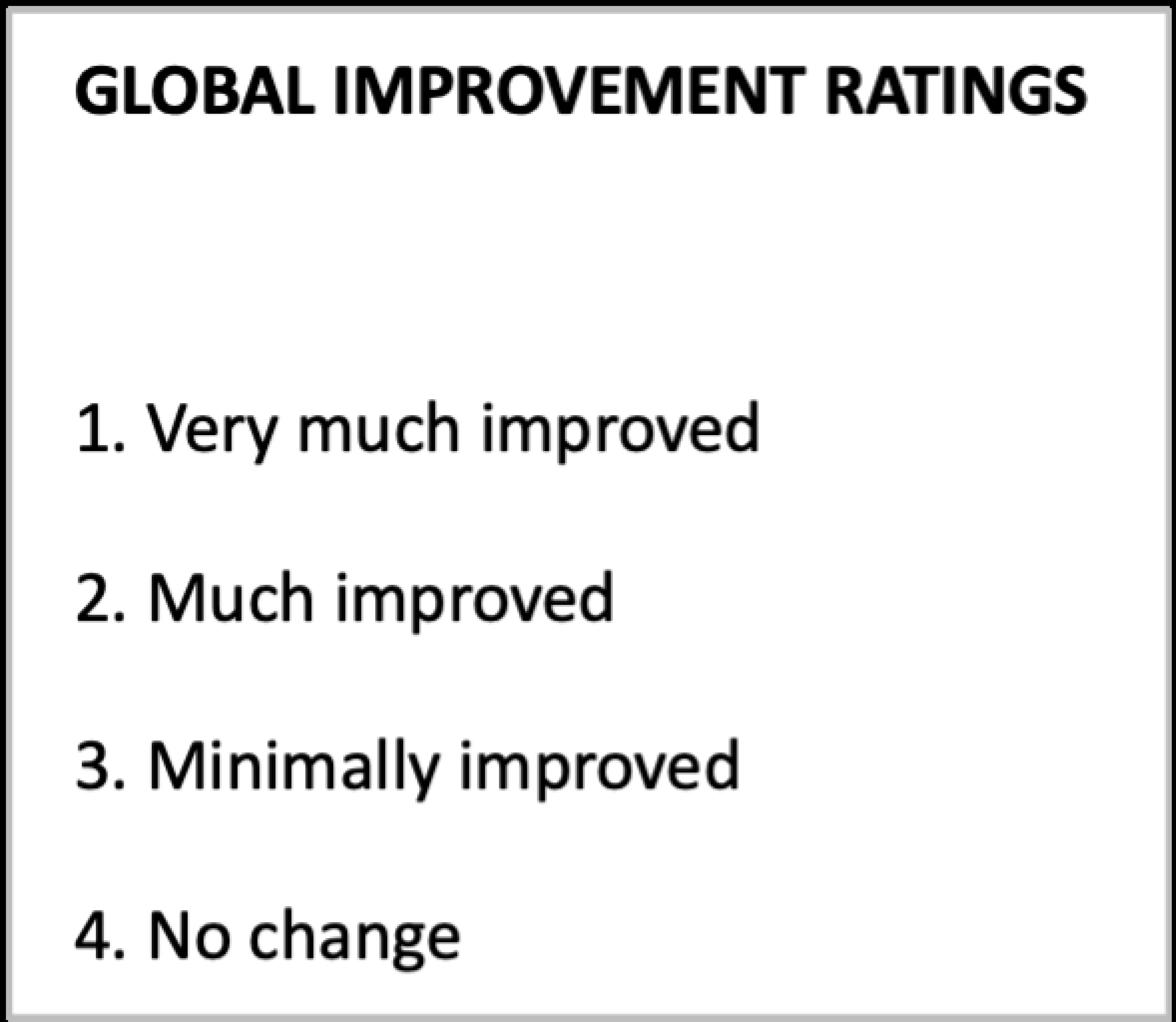
Clinical Global Impression – Severity of Illness.

At the end of the study, 14% of active patients receiving daily treatment with NTI-164 demonstrated the second highest possible efficacy index of 2, indicating marked therapeutic effects with side effects that did not significantly interfere with functioning. Clinicians rated NTI-164 to have a moderate therapeutic effect (efficacy index of 5 or 6) in 72% of treated patients, with half having no side effects and the other half having side effects that did not significantly interfere with functioning. A minimal therapeutic effect (efficacy index of 9) was observed in 7%, and an unchanged therapeutic effect (efficacy index of 13) in just one participant (Figure 3).

**Fig 3.**
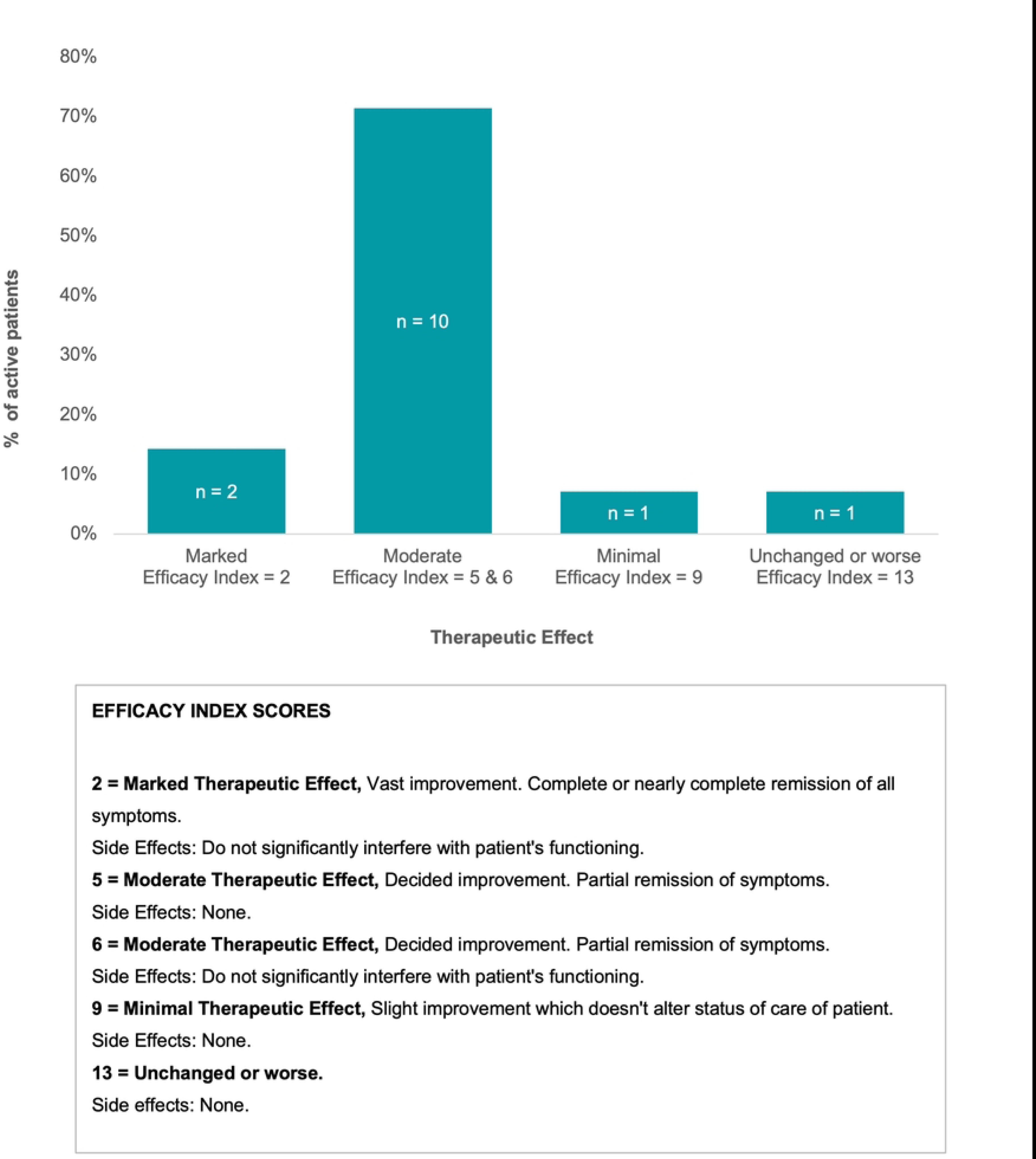
Clinical Global Impression - Therapeutic Effect after 28 days of treatment.

#### Anxiety symptoms

To measure anxiety symptoms in treated participants with ASD, the Anxiety Scale for Children, both -Parent and -Child versions (ASC-ASD) were used. The mean scores, Wilcoxon Signed-Rank test, and corresponding *p* values for both ASC-ASD-P and ASC-ASD-C versions are summarised and reported in Table 5. The total score of the Anxiety Scale for Children was reduced in more than half of both parents (62%) and children (54%) scales after four weeks of treatment. During the same period, nearly a quarter of parents (23%) and a third of children (36%) reported increased levels of anxiousness, while 15% and 9% of parents and children reported no change, respectively. The mean change for the total score of the ASC-ASD parent version was significantly reduced by 14% (*p*=0.0414), while the child version of the scale was non-significantly reduced by 6.8% after four weeks of treatment. The most frequently endorsed items on the ASC-ASD-P and ASC-ASD-C were from the Uncertainty (mean change 15.6%) and Separation Anxiety (25.4%) subscales post-treatment, respectively (Table 5).

**Table 5.** Anxiety assessment for Children. Results are expressed as Mean ± SD.

| Assessment | Baseline | Week 5 | Mean<br>change | Wilcoxon signed<br>rank test<br>($P$ value) |
| --- | --- | --- | --- | --- |
| *Anxiety Scale for Children – Parent Version (ASD-ASC-P) n=11* |  |  |  |  |
| Total ASC-ASD | 31.5± 14.7 | 27.1± 12.9 | -4.4 | 47.5 (0.0414) |
| Performance Anxiety | 7.5± 4.4 | 6.5± 3.7 | -1.0 | 25.5 (0.0474) |
| Anxiety Arousal | 4.0± 3.9 | 3.9± 3.2 | -0.1 | 15.0 (0.865) |
| Separation Anxiety | 5.3± 4.1 | 4.9± 4.0 | -0.4 | 17.5 (0.139) |
| Uncertainty | 14.1± 5.8 | 11.9± 4.4 | -2.2 | 47.0 (0.0459) |
| *Anxiety Scale for Children – Child Version (ASD-ASC-C) n=9* |  |  |  |  |
| Total ASC-ASD | 30.9± 10.1 | 28.8± 13.3 | -2.1 | 33.0 (0.213) |
| Performance Anxiety | 6.6± 3.9 | 6.1± 3.3 | -0.5 | 21.0 (0.206) |
| Anxiety Arousal | 4.3± 3.7 | 5.4± 3.1 | 1.1 | 11.5 (0.360) |
| Separation Anxiety | 6.7± 3.6 | 5.0± 4.4 | -1.7 | 37.0 (0.0834) |
| Uncertainty | 13.3± 4.9 | 12.2± 5.5 | -1.1 | 26.0 (0.258) |

#### Sleep quality and sleep disturbances

To assess changes to sleep quality, the SDSC questionnaire was used. Of the 14 study participants, 11 children completed sleep assessment before and after 28 days of treatment. Although the total sleep score decreased from baseline to post-treatment, it was not statistically significant. The scores for all sleep outcomes except Disorders of excessive somnolence improved non-significantly from baseline to post-treatment (Table 6).

**Table 6.** Sleep Disturbances Scale for Children (SDSC). Results are expressed as Mean ± SD.

| Sleep Domains | Baseline | Week 5 | Mean<br>difference | Wilcoxon<br>signed-rank<br>test<br>(p value) |
| --- | --- | --- | --- | --- |
| Total Score (n=11) | 50.5± 17.1 | 48.4± 13.4 | -2.0 | 29 (0.721) |
| Disorder of initiating &<br>maintaining sleep | 17.6± 6.9 | 16.4± 6.3 | -1.2 | 24.5 (0.811) |
| Sleep breathing disorders | 6.0 ± 3.0 | 5.5± 2.3 | -0.5 | 16 (0.234) |
| Disorders of arousal | 4.0± 1.6 | 3.9± 1.2 | -0.1 | 3 (1) |
| Sleep-wake transition disorders | 11.9± 4.4 | 11.2± 3.3 | -0.7 | 15 (0.343) |
| Disorders of excessive somnolence | 8.3± 3.3 | 9.1± 3.7 | 0.8 | 13 (0.481) |
| Sleep hyperhydrosis | 2.7± 1.2 | 2.4± 0.9 | -0.3 | 5 (0.285) |

## Discussion

The current pharmacological and behavioural interventions are limited in treating a considerable portion of children with ASD-related symptoms due to a lack of efficacy or adverse side effects that interfere with daily functioning. Therefore, there is an urgent need for a more concerted effort in clinical research to develop effective and safer therapies for addressing ASD associated comorbid symptoms. While the available evidence from controlled studies on the role of medicinal *Cannabis sativa* in the management of children with ASD is limited, a large body of evidence from preclinical studies has demonstrated the positive effects of cannabinoids in the modulation of a wide range of biological responses in particular neurodevelopmental processes. Furthermore, emerging evidence from recent observational studies [53–55, 57] has indicated that a synergistic and entourage pharmacological effect of *Cannabis sativa* derivatives can contribute to improved ASD-related symptoms and comorbidities in children. Therefore, this study sought evidence for the safety and efficacy of NTI-164, a full spectrum botanical product, per the entourage effect in managing ASD comorbid symptoms in paediatric patients. The findings demonstrate that NTI-164 is safe and well-tolerated in children up to 20 mg/kg daily dose with transient side effects that did not significantly interfere with the general functioning of treated patients.

Furthermore, NTI-164 exhibited statistically significant efficacy in improving ASD core and associated symptoms after four weeks of treatment. In particular, following NTI-164 treatment, targeted behavioural problems were much improved or very much improved in nearly half of the patients. More than half of caregivers and children also reported consistent improvements in the anxiety level of participants. Despite the limited sample size with completed sleep data, the overall direction of change indicated improved sleep outcomes and warrants further longitudinal evaluation. Based on these promising findings, we have initiated an extended treatment open-label trial for 57 weeks to assess long-term NTI-164 safety and efficacy in children and adolescents with ASD and related comorbidities.

### Impact of full-spectrum cannabis extract on ASD core symptoms

The safety profile and tolerability of NTI-164 were favourable in the patients who completed treatment consistent with previous clinical studies that used whole plant medicinal cannabis extract to treat children and adolescents with ASD-related symptoms [52–58]. The most frequent side-effect in our cohort was related to gastrointestinal symptoms, which were transient and improved with rest by the end of the treatment phase. However, the majority of clinical studies using full-spectrum cannabis extract for the treatment of children with ASD reported mild or transient adverse events frequently related to sleep disturbances [52–55, 58, 59], anxiety or irritability symptoms [52, 55, 57, 58] which were absent in our cohort. This might be due to different strains with variable composition of phytocannabinoids and higher doses of THC than our full-spectrum medical cannabis extract with negligible THC (0.08%). Alternatively, side-effects not observed in our study might be due to other studies, including primarily children diagnosed with severe ASD, since sleep disturbances have been reported as a side-effect of isolated CBD treatment in children with severe behavioural problems [68]. In addition, several studies have found that the severity of ASD core symptoms is associated with the severity of sleep disturbances in younger children [60, 69, 70]. Compared to other clinical studies that included children as young as four years old, our active patients were much older (10-17 years). The effect of different compositions of phytocannabinoids and THC levels in full-spectrum extracts on the severity of ASD symptoms in younger children, therefore, requires further investigation.

This is the first study of full-spectrum cannabis extract treatment in children with ASD to report subjective measures of adverse events and objective measures of biochemical analysis and vital signs following treatment. In a prospective cohort study of biochemical safety analysis of an oil-based full-spectrum cannabis product, all parameters were within the normal range in children with ASD after three months of treatment [71]. However, this study did not report an evaluation of adverse events or vital signs following treatment. Notably, a recent randomised controlled feasibility study reported no significant abnormal biochemical test results after eight weeks of therapy with isolated CBD in children with severe behavioural problems and intellectual disability [68].

In our study, overall daily treatment with NTI-164 for four weeks significantly improved symptoms in 93% of active patients, with nearly two-thirds of children rated by clinicians to exhibit moderate effects following treatment. Similarly, several cohort studies in children using full-spectrum products have demonstrated significant overall improvement in ASD symptoms reported by parents [52–54, 56] and clinicians [55, 57]. Specifically, we found that almost half of treated children with ASD showed reduced disruptive behaviour consistent with other studies that have demonstrated improvement in behavioural problems ranging from 32% to 90%, including adaptive behaviours, self-injury and hyperactivity [53, 57, 59]. Social interaction and communication have also significantly improved following treatment with full-spectrum cannabis extract in children with ASD-related symptoms [52, 55–57]. In addition, two randomised controlled trials have shown that cannabinoid treatment, compared to placebo in children, was safe with acceptable tolerability and effective at reducing ASD-associated disruptive behaviours [58, 59]. However, compared to pure cannabinoids, there was no apparent difference with improvements observed for treatment with whole-plant extract after 12 weeks. The clinical benefits of the entourage effects of our cannabis strain compared to purified cannabinoids remain to be further explored in longitudinal controlled studies.

### Impact of full-spectrum cannabis extract on ASD-related comorbidities

In addition to improvements in behavioural outcomes, a recent systematic review of the evidence for medicinal cannabis in ASD treatment found benefits for a broad range of comorbidities, including cognition, anxiety and sleep disturbances in adults and children [72]. Consistent with previous studies [52–54], we also found that after four weeks of treatment with full-spectrum cannabis extract, anxiety symptoms were reduced in more than half of our cohort based on both parent and children reported anxiety questionnaires. In particular, the parent-rated total scores and the performance anxiety and uncertainty subscales significantly improved for children with ASD. However, the magnitude of improvement on child-rated outcome measures did not reach statistical significance despite the direction of change, suggesting reduced anxiety symptoms in half of the treated patients. This rating discrepancy might be due to differences in perceptions of anxiety for children with ASD and their parents with the severity of ASD symptoms reportedly contribute to the disparity in parent-child anxiety rating where higher severity levels were shown to be associated with the tendency for parent ratings to exceed child anxiety rating [73]. Nevertheless, this finding highlights the clinical implication of self-report rating to inform areas of concern for continued assessment of anxiety symptoms in children with ASD.

While previous prospective cohort studies have reported treatment with whole plant cannabis extract of various strains significantly improved sleep outcomes in children with ASD [53–55], a recent randomised controlled trial did not find any difference between cannabinoid treatment and placebo in sleep parameters after 12 weeks [60]. We also did not find significant improvement in any of the sleep domains as measured by a validated sleep questionnaire. These inconsistencies could be due to differences in treated participant characteristics, especially level of ASD severity, age, instruments used to measure sleep or strain of cannabis with various phytocannabinoid composition. In addition, since the previous studies that reported improved sleep outcomes were over several months, this may indicate the need for a longer period before any benefit in sleep quality can be observed from treatment with cannabinoid extract.

These findings support the safety and efficacy of full-spectrum products and promising entourage effects of cannabinoids in treating children with ASD-related symptoms.

### Limitations

There were several limitations in this study. Since our study was an open-label design to collect preliminary data on the safety and efficacy of NTI-164 treatment of symptoms in children with ASD, no control group was included. However, the addition of objective measures of vital signs and biochemical analysis to subjective reports of adverse events further strengthened the NTI-164 safety profile in treated children with ASD. Nevertheless, further research for long-term evaluation of safety data of NTI-164 botanical product for the treatment of children with ASD symptoms, especially in terms of interaction with concomitant medications, is needed.

Given that the sample size was small, evaluation of the impact of NTI-164 treatment on subgroups of children with various ASD severity was not possible. Although data was collected on concomitant medications prescribed for ASD-related comorbidities, this study was not powered to determine the impact of chronic medication on treatment response. Also, we only used subjective measures of sleep outcomes, and future studies should include actigraphy or polysomnography together with sleep diaries as objective measurements of sleep quality.

## Conclusions

This open-label study provided preliminary efficacy and safety data on a novel botanical formulation of phytocannabinoids in treating symptoms in children with ASD and associated comorbidities. Results indicate that NTI-164 is safe and well-tolerated, with significant efficacy in improving disruptive behaviours and reducing anxiety in this Pediatric cohort. The findings of this study further contribute to the accumulating evidence in favour of whole plant cannabis extracts, with most participants reporting a clinically meaningful change in ASD-related symptoms and associated anxiety. Despite the small sample size, objective safety analysis together with converged evidence from standardised clinician, parent and child-reported questionnaires with respect to overall improvement strengthen the evidence for the benefit of full-spectrum cannabis extract in the treatment of children with ASD-related symptoms. Furthermore, these results are consistent with other prospective cohort studies of full-spectrum cannabis extract and support further evaluation of the clinical benefits of whole plant extracts in rigorous placebo-controlled trials of longer duration in this patient population.

## Data Availability

All relevant data are within the manuscript and its Supporting Information files.

## Notes

**Funding** This study was supported by Fenix Innovations.

### Competing Interest Statement

I have read the journal's policy and the authors of this manuscript have the following competing interests: The study was funded by Fenix Innovations.

### Clinical Trial

This study has been registered with the Australian and New Zealand Clinical Trial Registry (Trial ID: ACTRN12621000760875)

### Funding Statement

Yes

### Author Declarations

This study was approved by the Monash Health Human Research Ethics Committee - RES-21-0000-177A

